# A phase III double-blind, placebo-controlled, randomized withdrawal trial of 5-aminolevulinic acid hydrochloride with sodium ferrous citrate for efficacy and safety in patients diagnosed as Leigh syndrome

**DOI:** 10.1101/2025.08.31.25334800

**Authors:** Akira Ohtake, Yuichi Abe, Kei Murayama, Hideaki Shiraishi, Hitoshi Osaka, Satoko Kumada, Tetsuya Ito, Hidehito Kondo, Yusuke Hamada, Sayaka Ajihara, Masato Arao, Yuki Ueda, Kohei Nagai, Keiko Ichimoto, Ayako Matsunaga, Hirofumi Kashii, Yoko Nakajima, Hitoshi Nakagawa, Shunichi Kurose, Kaori Fukuda, Kiwamu Takahashi, Mitsugu Yamauchi, Motowo Nakajima

## Abstract

**Objective:** A phase III, double-blind, placebo-controlled, randomized withdrawal trial of SPP-004 (5-aminolevulinic acid hydrochloride and sodium ferrous citrate) in patients diagnosed as Leigh syndrome (LS) was conducted to confirm the efficacy and safety of SPP-004 in patients with LS showing central nervous system disorders.

**Methods:** Fifty-four patients entered a 24-week open-label period of SPP-004 administration. Among them, 28 patients showing improved scores on the Newcastle Paediatric Mitochondrial Disease Scale (NPMDS) for cranial nervous symptoms and myopathy symptoms proceeded to a 48-week double-blind (DB) period, where they were randomized (1:1) to receive SPP-004 or placebo (*n*=14 each). Efficacy was evaluated using NPMDS for the full analysis set (FAS) during the DB-period (SPP-004 *n*=13, Placebo *n*=14) and the entire study period (*n*=54). Safety evaluation focused on adverse events (AEs) in all 54 patients administered SPP-004.

**Results:** The primary endpoint, the proportion of patients who discontinued due to inadequate efficacy at 48 weeks, was lower in the SPP-004 group (15.4% [95% CI: 1.9-45.4%]) compared to the placebo group (50.0% [23.0-77.0%]). Over 80% of the SPP-004 group maintained efficacy (p=0.0486). All adverse drug reactions were mild, with no notable differences in AEs between groups.

**Conclusion:** These findings suggest that SPP-004 is safe and may provide therapeutic effect for LS symptoms.

## Introduction

Mitochondrial diseases (MD) are clinically and genetically heterogeneous disorders caused by mutations in genes encoded in the nuclear DNA or by mutations and/or deletions in the mitochondrial DNA. MD is associated with progressive metabolic disorders caused by dysfunctional energy production in the mitochondrial respiratory chain (MRC) [1,2]. MD is also referred to as mitochondrial respiratory chain disorder (MRCD) [3]. Patients with MD often develop hyperlactatemia, diagnosed by elevated lactate levels in both blood and cerebrospinal fluid, which often affects the central nervous system and various systemic organs [1].

Leigh syndrome (LS) is a common type of MD characterized by central nervous system abnormalities, including bilateral basal ganglia lesions in infancy, detectable by brain magnetic resonance imaging (MRI) [4]. Patients with LS experience psychomotor delays in infancy or childhood and progressively develop neurological regressions. LS is confirmed through genetic examination for mitochondrial and nuclear DNA mutations [5,6]. Mitochondrial functions in patients with MD, including those with LS and MRC complex I deficiency, can be evaluated through biochemical examination of MRC complex enzyme activity and oxygen consumption using fibroblasts from a primary culture of a skin biopsy sample [7].

Despite the severity of MD, the patients with MD, including LS and mitochondrial myopathy, encephalopathy, lactic acidosis, and stroke-like episodes (MELAS) are treated with oral multivitamin therapy, nutrition therapy with a lipid diet, and supportive therapies [8]. Magro et al. [9] have recently published a comprehensive review on LS; disease and present and future treatments, which nicely summarizes therapeutic and supplemental materials for LS patients. Although several clinical trials of MD therapeutics, including orally available vatiquinone (EPI-743, PTC743) for inherited MDs such as MELAS, LS, Friedreich ataxia (FA), and Alpers syndrome, have been conducted globally [10–13], only a few drugs have been approved for specific MD symptoms [9,10].

5-aminolevulinic acid (5-ALA), combined with sodium ferrous citrate (SFC), has been reported to enhance heme production [14] and upregulate MRC activity *in vitro* [15]. Oral administration of 5-ALA was observed to facilitate brain mitochondrial activity in a mouse model of Alzheimer’s disease [16]. Furthermore, in a rat model of autism spectrum disease (ASD), oral administration of 5-ALA ameliorated oxidative stress and mitochondrial dysfunction and increased ATP production in the hippocampus, and 5-ALA administration improved ASD-like neuropathology [17].

Nozawa et al. [18] recently reported that a combination of 5-ALA hydrochloride (5_-_ALA-HCl) and SFC increased ATP production and suppressed defective phenotypes in *Drosophila* with MRC CI deficiency. Knockdown of *sicily*, a *Drosophila* homolog of the Complex I assembly protein NDUFAF6, caused Complex I deficiency, lactate and pyruvate accumulation, and detrimental phenotypes such as abnormal neuromuscular junction development, locomotor dysfunction, and premature death. Feeding with 5-ALA-HCl and SFC increased ATP levels without recovery of Complex I activity, while the activities of Complexes II and IV were upregulated, and lactate and pyruvate accumulation was suppressed. Feeding of 5-ALA-HCl and SFC improved neuromuscular junction development and locomotor functions in *sicily*-knockdown flies. These results suggest that supplementation with 5-ALA-HCl and SFC shifts metabolic programs to address mitochondrial Complex I deficiency, a major cause of LS.

Since no drug was available for LS therapy, we conducted an investigator-initiated trial to evaluate the efficacy and safety of SPP-004 in patients with LS in comparison with placebo administration. Preliminary safety and efficacy of 5-ALA-HCl and SFC in pediatric patients with LS were suggested by results from a 24-week exploratory trial, followed by continuous long-term administration of SPP-004 for up to 204 weeks [19]. Since very few pediatric patients with LS were available in the first double-blind trial, a subsequent trial with a large cohort of patients with LS was necessary to confirm the safety and efficacy of SPP-004. We designed a confirmatory trial with a longer double-blind period (48 weeks), which is the double-blind, placebo-controlled, randomized withdrawal trial of SPP-004 by recruiting Japanese patients with LS (aged 3-months or older), and the total score of Newcastle Paediatric Mitochondrial Disease Scale (NPMDS) for Sections I and III items 3–8 was employed as the primary endpoint, because the NPMDS Sections II and III items 1-2 are unsuitable for short-term efficacy evaluation [20].

Our previous study suggested that blood fibroblast growth factor 21 (FGF21) level might be a predictable surrogate marker for disease severity [19], as others have reported [21–24]. Blood growth differentiation factor 15 (GDF15) has also been reported to be a potential biomarker for mitochondrial dysfunction [24–26]. Therefore, we monitored both blood FGF21 and GDF15 levels.

## Methods

### Study design

The design of this study is shown in Fig 1A. This phase III investigator-initiated multicenter, randomized withdrawal, placebo-controlled, double-blind, parallel-group, confirmatory study (SPED-ALA-003 study) was conducted from July 2018 to May 2020 at eight institutions, including Saitama Medical School Hospital, Hokkaido University Hospital, Jichi Medical University Hospital, Chiba Children’s Hospital, Tokyo Metropolitan Neurological Hospital, Fujita Health University Hospital, Japanese Red Cross Society Kyoto Daiichi Hospital and Toyonaka Municipal Hospital with clinical trial number of jRCT2091220358.

**Fig 1.**
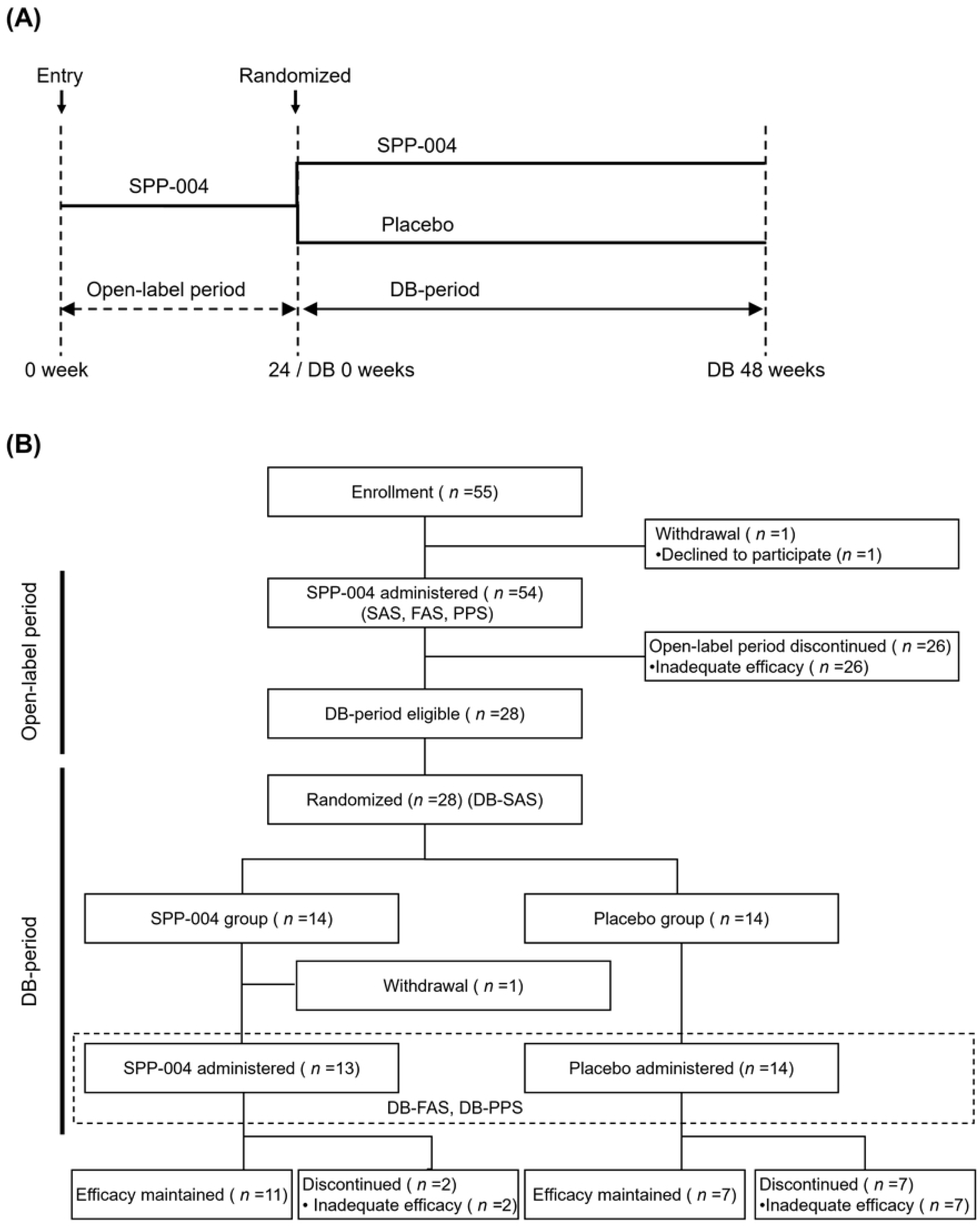
Study design (A) and Flow of subject disposition (B) DB-SAS: Safety analysis set (SAS) in the double blind (DB)-period, DB-FAS: Full analysis set (FAS) in the DB-period, DB-PPS: Per protocol set (PPS) in the DB-period

Because LS is a severe disease with poor prognosis, conducting a traditional placebo-controlled, double-blind study may cause ethical concerns that patients in the placebo group remain untreated for a long period. This study was conducted as a randomized withdrawal study that could minimize the duration of an untreated placebo to avoid such ethical risk. At the start of the study, follow-up observations were to be performed after Week 48 of the period or when treatment was discontinued. However, it was later changed to be optional for patients who wish for early entry into subsequent long-term administration study (SPED-ALA-004 study: jRCT2091220406). The protocol of this phase III study was reviewed and approved by the institutional review board of each study site as well as the relevant regulatory agency, and it was conducted in compliance with good clinical practice (GCP). The quality and reliability of this clinical trial were controlled by “quality control of the clinical trial” and “quality assurance of the clinical trial,” and the auditor guaranteed that the study was conducted in accordance with the study protocol and GCP. This study was registered at the <u>JRCT registry</u> with Trial ID: jRCT2091220358.

### Patients

Japanese patients with LS who were mainly involved in central nervous system disorders consented to participate and were judged to be eligible. They were enrolled for this investigator-initiated clinical study. Eligibility for this study and transition criteria for double-blind (DB) period are shown in Table 1.

**Table 1.**
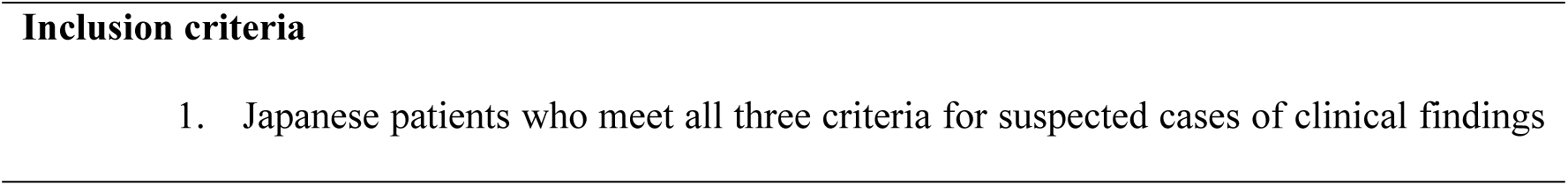
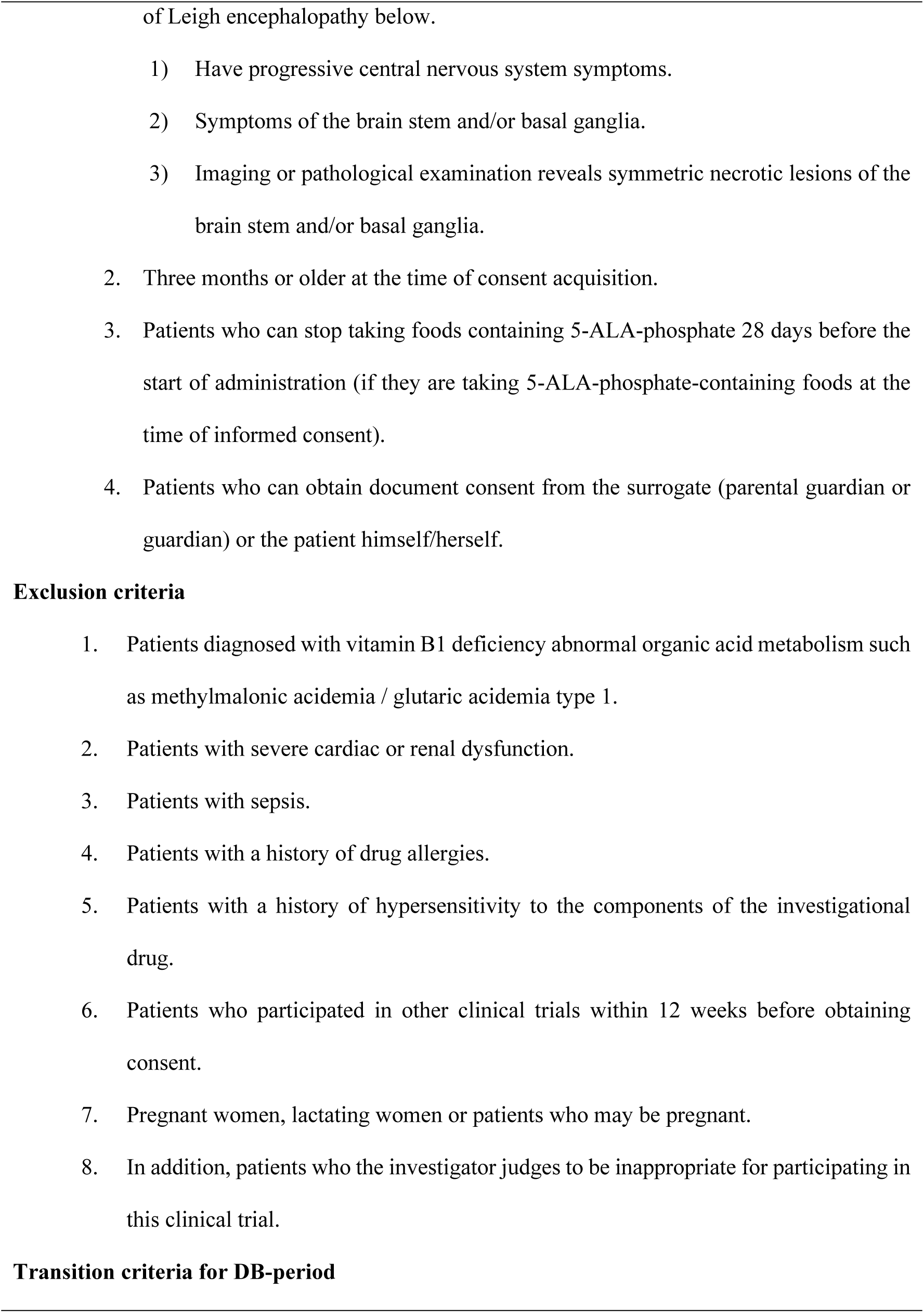
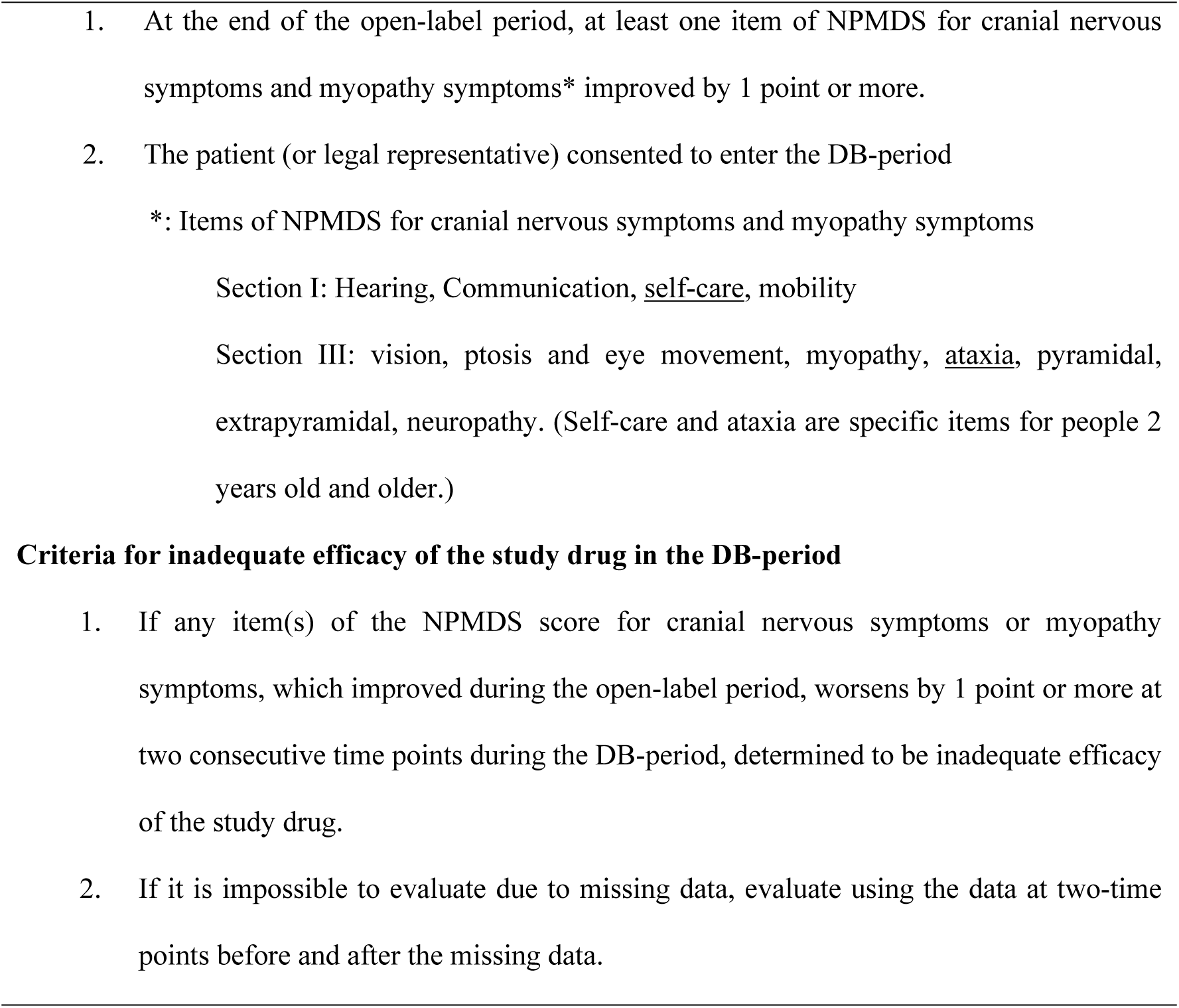
Criteria for Inclusion, exclusion, DB-period transition, and inadequate efficacy of the study drug.

During the open-label period, the patients were administered SPP-004 for 24 weeks. At the end of the open-label period, the patients who met the transition criteria entered the DB-period. Then, the DB-period transition patients were randomly assigned in a 1:1 ratio via electronic data capture according to randomized key cord and received SPP-004 or placebo for 48 weeks, respectively. The randomization key code was created by the allocation manager using computer-generated pseudo-random numbers based on a permuted block method with institution as the blocking factor.

A target size was set using the confidence interval method. At the time of planning the maximum number of patients that could be enrolled was 40, and assuming that 30 would proceed to the DB-period, and that 20% (3 patients) of the 15 cases in the SPP-004 group and 60% (9 patients) of the 15 cases in the placebo group would be inadequate efficacy, the exact 95% confidence interval (CI) for the difference in the rate of inadequate efficacy was calculated to be 1.2% to 70.5%, with the lower limit of the 95%CI exceeding 0. Based on these results, the target number of enrollment was set at 40, to ensure 30 patients eligible for randomization.

Discontinuation criteria requested for cancellation, obviously ineligible cases, cases with inadequate efficacy of the study drug during the DB-period, cases that are difficult to continue the trial due to adverse event (AE), worsening of underlying disease or decision of discontinuation by the investigator or sub-investigator.

Concomitant medications (5-ALA-HCl-containing a drug, iron-containing medicines including iron-based supplements, an investigational drug for mitochondrial disease conducted for other trials, and foods containing 5-ALA-phosphate, etc.) and concomitant treatments were prohibited throughout the study period to avoid interference for efficacy.

### Study drug and administration

The active drug, SPP-004, is a combination of SPP-004A capsules containing 25 mg of 5-ALA-HCl and SPP-004S capsules containing 39 mg of SFC. Placebo is a combination of capsules indistinguishable from SPP-004A without 5-ALA-HCl and capsules that are indistinguishable from SPP-004S containing 20 mg SFC. During the open-label period, the patients received SPP-004 orally or by tube at the doses shown in S1 Table, which were comparable to the doses that were administered in the exploratory trial [19]. During the DB-period, the patients received the same number of capsules of SPP-004 or Placebo, depending on the treatment group. During the DB-period, the study was conducted under blind conditions for all investigators (subjects, investigators, etc.) other than the allocation manager of the study drug.

### Efficacy and safety

The main target analysis population for efficacy in the DB-period was the largest analysis target population (full analysis set: FAS) based on the principle of intention-to-treat (ITT). Auxiliary analysis was performed on the population that fits the study protocol (per protocol set: PPS). The safety analysis was performed on the safety analysis set (SAS). The primary endpoint of the efficacy of this study was the proportion of patients who discontinued due to inadequate efficacy of the study drug at 48 weeks in the DB-period and their 95% CI (FAS of the DB-period). The exact 95% CI for the difference in proportions between the groups and the difference in proportions was also calculated. As a stratification analysis using the age at onset (under 1 year old, 1 year old, or older) of the primary disease as a stratification factor, the difference in proportions between groups was compared by the Mantel-Haenszel method. The 95% CI of the difference of proportion and its p-value were also calculated. The criteria for inadequate efficacy of the study drug in the DB-period is shown in Table 1.

The secondary endpoints of this study were the proportions of patients who discontinued due to inadequate efficacy of the study drug at 24 weeks and 36 weeks in the DB-period and a Kaplan-Meier plot for the duration of the DB-period until discontinuation that was calculated for each group regarding “inadequate efficacy of the study drug” as an event. A log-rank test was performed as a comparison between the two groups, and the p-value was calculated. Similar analyses were also planned to be performed for prognosis regarding death as an event but were not performed because no patient was dead in this study.

In this study, the Newcastle Pediatric Mitochondrial Disease Scale (NPMDS) [20] was mainly used for the evaluation of the efficacy of investigational drugs. There are three versions of NPMDS (for 0-24 months, 2-11 years, 12-18 years) depending on the age group. Since the purpose of this trial was to evaluate changes in scores, even if the age group was changed during the course, the evaluation was performed using the NPMDS version of the age at the time of enrollment during the open-label period.

For NPMDS, the total score (cranial nervous symptoms and myopathy symptoms, Section I to III, Section I to IV, each section) and the summary statistics of the measured values, the changes from baseline at each evaluation point for each item were calculated for each group (open-label period withdrawal, SPP-004 and Placebo). The items for the cranial nervous symptoms and myopathy symptoms included hearing, communication, self-care, and mobility in section I, vision, ptosis, and eye movement, myopathy, ataxia, pyramidal, extrapyramidal, and neuropathy in section III. Self-care and ataxia were specific items for 2 years old and older.

For SPP-004 and Placebo groups, the change from the start of the DB-period was also evaluated. The analysis using the data complemented by the LOCF (Last observation carried forward) method was also performed in the same manner. For FAS in the DB-period, A Swimmers Plot, which shows the period from the start of the DB-period to discontinuation / the end of the study for each case, was created. Heat maps were created to show changes in scores for cranial nervous symptoms and myopathy symptoms in each patient. Open-label efficacy was defined as an improvement in the score at 24 weeks of the open-label period compared to the baseline score. Long-term efficacy was defined as the open-label efficacy maintained for up to 44 weeks of the DB-period, and short-term efficacy was defined as the open-label efficacy maintained for less than 44 weeks of the DB-period.

Outcomes of efficacy other than NPMDS were blood lactate, blood FGF21, blood GDF15, body weight, height, the enzyme activity of respiratory chain complex and oxygen consumption rate in dermal fibroblasts, brain MRI change, prognosis, overall improvement, the period without ventilator and number of stroke-like seizures (only for subjects with stroke-like seizure symptoms before the start of the trial).

Blood lactate was measured by the clinical laboratory of each hospital as routine blood analysis. Blood FGF21 was measured by using ELISA kits for FGF21 (BioVendor, Brno, Czech Republic) in Dr. Otake’s laboratory at Saitama Medical University. Blood GDF15 was measured by LSI Medience (Tokyo, Japan).

For safety assessment, terms of AEs described by the doctors were encoded by using Medical Dictionary for Regulatory Activities (MedDRA) /J Ver 23.0 and classified by the group, study period, relevance to the study drug, severity, system organ class (SOC), and preferred term (PT).

For laboratory test values and vital signs, summary statistics at each observation point were calculated for each group. Subject frequencies and proportions were calculated for categorical variables, and descriptive statistics were calculated for continuous variables.

### Statistical analysis

The Windows version of SAS release 9.4 or higher was used for statistical analysis. Unless otherwise noted, the test was a two-sided hypothesis with a significance level of 5%. The confidence interval for the confidence coefficient was 95% on both sides. The confidence interval for the ratio was calculated using the Clopper-Pearson exact method. If necessary, analyses using data complemented by the Last Observation Carry Forward (LOCF) method were also performed to assess the effects of missing values due to early discontinuation.

## Results

### Patient flow and demographics

The study design and the patient flow diagram are shown in Fig 1A and 1B, respectively. From July 25, 2018 to May 26, 2020, 54 of 55 Japanese patients with LS who met eligibility criteria and enrolled in the study entered the open-label period for administration of SPP-004, while one patient was excluded due to the investigator’s decision. Twenty-six patients dropped out during the 24-week open-label period. The other 28 patients, who showed improvement in at least one item of cranial nervous symptoms and myopathy symptoms, entered the DB-period, and 14 patients each were assigned to the SPP-004 and the placebo groups, respectively. Efficacy for DB-period was assessed in 27 patients (FAS of DB-period and PPS of DB-period), including 13 in the SPP-004 group and 14 in the placebo group, excluding one with no efficacy data of the DB-period in the SPP-004 group.

Table 2 shows the demographic and other baseline characteristics of the patients of FAS. Regarding gender, the male-female ratios were similar in the open-label period, 55.6% (30/54) for males and 44.4% (24/54) for females. However, there was a gender bias between the two groups in the DB-period; 38.5% (5/13) males and 61.5% (8/13) females were assigned to the SPP-004 group, whereas 85.7% (12/14) males and 14.3% (2/14) females were allocated to the placebo group.

**Table 2.** Demographic and other baseline characteristics (FAS)

|  | *n* (%) |  |  |  |  |
| --- | --- | --- | --- | --- | --- |
|  | Open-label period |  |  | DB-period |  |
|  | Total | discontinued | Complete | SPP-004 | Placebo |
| **Patients** | 54 | 26 | 28 | 13 | 14 |
| **Sex** |  |  |  |  |  |
| Male | 30 (55.6) | 12 (46.2) | 18 (64.3) | 5 (38.5) | 12 (85.7) |
| Female | 24 (44.4) | 14 (53.8) | 10 (35.7) | 8 (61.5) | 2 (14.3) |
| **Age at the time of informed consent** |  |  |  |  |  |
| Mean ± S.D. | 7.9 ± 6.52 | 8.3 ± 6.86 | 7.5 ± 6.29 | 7.2 ± 6.30 | 7.4 ± 6.50 |
| Median [Max, Min] | 6.5 [1, 33] | 7.0 [1, 33] | 4.5 [1, 25] | 4 [1, 19] | 5.5 [1, 25] |
| < 2 years old | 4.0 (7.4) | 2.0 (7.7) | 2.0 (7.1) | 1.0 (7.7) | 1.0 (7.1) |
| >2 - ≤11 years old | 36 (66.7) | 16 (61.5) | 20 (71.4) | 9 (69.2) | 11 (78.6) |
| ≥ 12 years old | 14 (25.9) | 8 (30.8) | 6 (21.4) | 3 (23.1) | 2 (14.3) |
| **Primary disease** |  |  |  |  |  |
| Leigh | 48 (88.9) | 23 (88.5) | 25 (89.3) | 11 (84.6) | 13 (92.9) |
| Leigh, MELAS | 6 (11.1) | 3 (11.5) | 3 (10.7) | 2 (15.4) | 1 (7.1) |
| **Age of onset** |  |  |  |  |  |
| Mean ± S.D. | 1.7 ±2.74 | 1.6 ±2.52 | 1.8 ± 2.98 | 1.5 ±3.43 | 2.1 ±2.71 |
| Median [Max, Min] | 1 [0, 12] | 0 [0, 9] | 1 [0, 12] | 0 [0, 12] | 1 [0, 9] |
| < 1 year old | 26 (48.1) | 14 (53.8) | 12 (42.9) | 8 (61.5) | 4 (28.6) |
| ≥1 year old | 28 (51.9) | 12 (46.2) | 16 (57.1) | 5 (38.5) | 10 (71.4) |
| **Family history** |  |  |  |  |  |
| None | 36 (66.7) | 18 (69.2) | 18 (64.3) | 8 (61.5) | 10 (71.4) |
| Yes | 18 (33.3) | 8 (30.8) | 10 (35.7) | 5 (38.5) | 4 (28.6) |
| Medical history |  |  |  |  |  |
| None | 29 (53.7) | 18 (69.2) | 11 (39.3) | 6 (46.2) | 4 (28.6) |
| Yes | 25 (46.3) | 8 (30.8) | 17 (60.7) | 7 (53.8) | 10 (71.4) |
| Presence of complications |  |  |  |  |  |
| None | 5 (9.3) | 0 (0.0) | 5 (17.9) | 2 (15.4) | 3 (21.4) |
| Yes | 49 (90.7) | 26 (100.0) | 23 (82.1) | 11 (84.6) | 11 (78.6) |

The ages at the time of informed consent obtained were similar in the open-label period total (7.9 ± 6.52 years) and the DB-period; the SPP-004 group (7.2 ± 6.30 years) and the placebo group (7.4 ± 6.50 years). Regarding primary disease type, all patients had LS. Concurrent MELAS occurred in 11.1% (6/54) of all patients in the open-label period, 15.4% (2/13) in the SPP-004 group in the DB-period, and 7.1% (1/14) in the placebo group. Other underlying diseases were Kearns-Sayre syndrome and chronic progressive external ophthalmoplegia in one patient each, both of which were discontinued during the open-label period due to lack of efficacy.

The ages at onset of the primary disease were almost the same in the open-label period, with 48.1% (26/54) of those less than 1 year old and 51.9% (28/54) of those who were 1 year or older. However, during the DB-period, there were more children <1 year (61.5%: 8/13) than ≥1 year (38.5%: 5/13) in the SPP-004 group, whereas there were more children ≥1 (71.4%: 10/14) year than <1 year (28.6%: 4/14) in the placebo group. There were no group differences in family history, medical history, or complications. In conclusion, there was a difference between the groups in terms of gender, but due to the small number of cases, a detailed analysis was not performed.

### Efficacy results

Table 3 shows the primary endpoint of this study, the proportion of patients who discontinued the study drug at 48 weeks in the DB-period due to inadequate efficacy of the study drug (FAS of the DB-period). At 48 weeks of the DB-period, the proportion of patients with inadequate efficacy of the study drug was 15.4% (2/13 patients, 95% confidence interval [95% CI]:1.9% to 45.4%) in the SPP-004 group, which was lower than 50.0% (7/14 patients, 95% CI: 23.0% to 77.0%,) in the placebo group (difference in proportion −34.6% [95% CI: −65.7% to 2.4%]). The efficacy was more sustained in the SPP-004 group, but the difference was not significant (unadjusted analysis, p=0.1032). When adjusted by the Mantel-Haenszel method with age at onset of primary disease as a stratification factor (<1 year old, 1-year-old or older), the difference of the proportion of subjects judged to be “inadequate efficacy of study drug” between the SPP-004 group and the placebo group was 22.2% (95% CI: −56.9% to 12.4%), suggesting that efficacy of study drug was more sustained in the SPP-004 group, but the difference was not significant (adjusted analysis, p=0.2059). The same result was obtained for PPS of DB-period transitioned patients. Regarding the secondary endpoints, the proportion of subjects with inadequate efficacy of the study drug at weeks 24 and 36 in the DB period was the same as the results at 48 weeks in the DB-period.

**Table 3.**
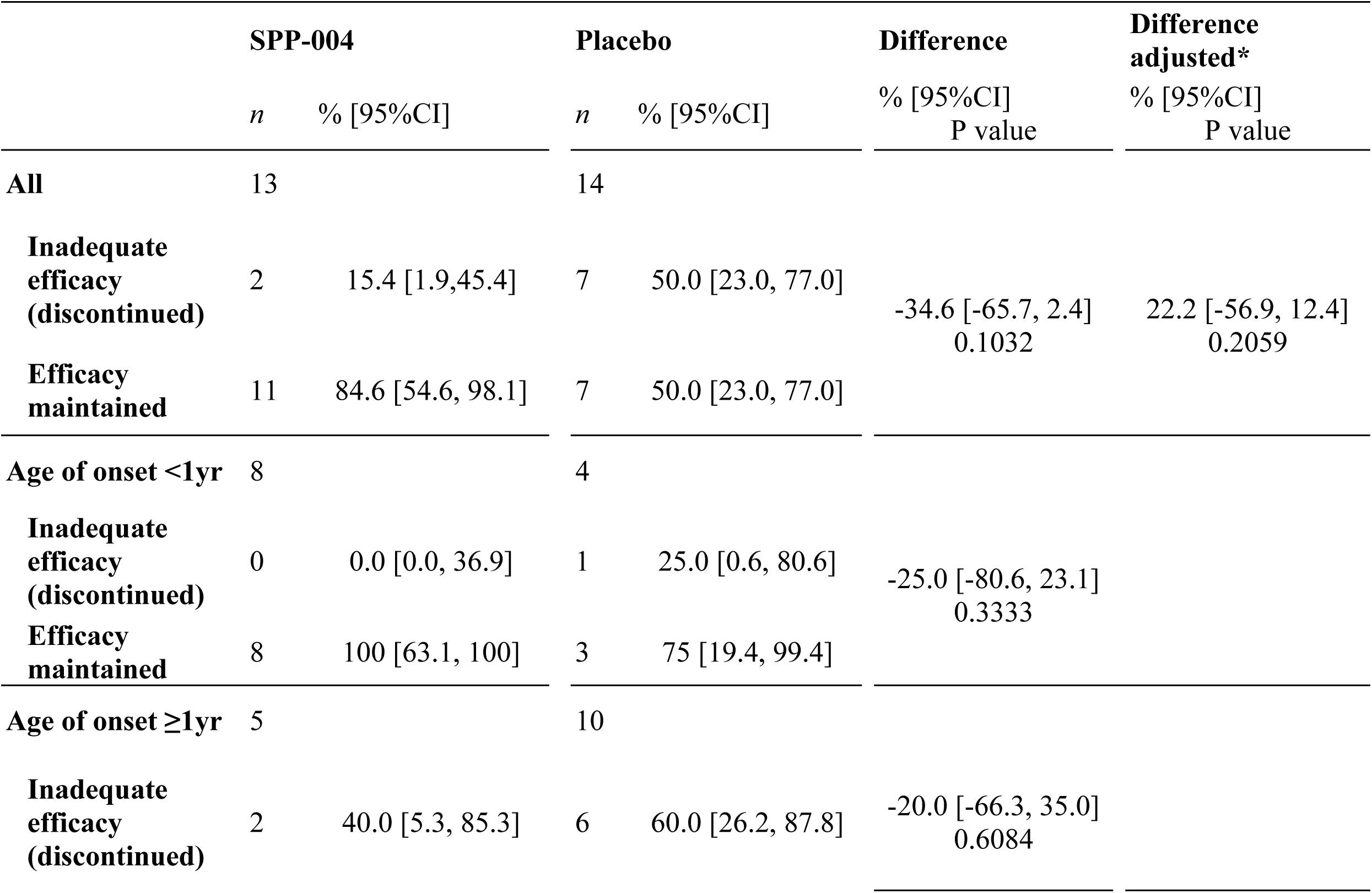
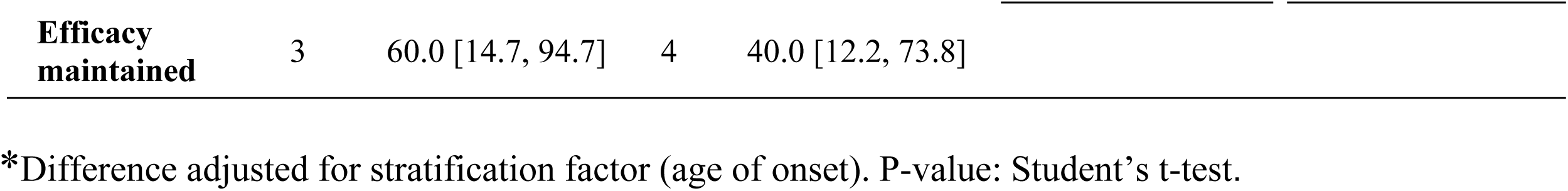
Proportion of patients with inadequate efficacy of the study drug at the 48 weeks in the DB-period as the primary endpoint (FAS in the DB-period)

|  | SPP-004 |  | Placebo |  | Difference | Difference adjusted* |
| --- | --- | --- | --- | --- | --- | --- |
|  | *n* | % [95%CI] | *n* | % [95%CI] | % [95%CI]<br>P value | % [95%CI]<br>P value |
| **All** | 13 |  | 14 |  |  |  |
| **Inadequate efficacy (discontinued)** | 2 | 15.4 [1.9,45.4] | 7 | 50.0 [23.0, 77.0] | -34.6 [-65.7, 2.4]<br>0.1032 | 22.2 [-56.9, 12.4]<br>0.2059 |
| **Efficacy maintained** | 11 | 84.6 [54.6, 98.1] | 7 | 50.0 [23.0, 77.0] |  |  |
| **Age of onset <1yr** | 8 |  | 4 |  |  |  |
| **Inadequate efficacy (discontinued)** | 0 | 0.0 [0.0, 36.9] | 1 | 25.0 [0.6, 80.6] | -25.0 [-80.6, 23.1]<br>0.3333 |  |
| **Efficacy maintained** | 8 | 100 [63.1, 100] | 3 | 75 [19.4, 99.4] |  |  |
| **Age of onset ≥1yr** | 5 |  | 10 |  |  |  |
| **Inadequate efficacy (discontinued)** | 2 | 40.0 [5.3, 85.3] | 6 | 60.0 [26.2, 87.8] | -20.0 [-66.3, 35.0]<br>0.6084 |  |

|  |  |  |  |  |
| --- | --- | --- | --- | --- |
| **Efficacy maintained** | 3 | 60.0 [14.7, 94.7] | 4 | 40.0 [12.2, 73.8] |

\*Difference adjusted for stratification factor (age of onset). P-value: Student's t-test.

Fig 2A shows the Kaplan-Meier curve of the period until discontinuation due to “inadequate efficacy of the study drug” in DB-period transitioned patients. In the Kaplan-Meier plot by treatment group with discontinuation due to inadequate efficacy of the study drug as an event, when censored at 48 weeks of the DB-period, more than 80% of the patients in the SPP-004 group maintained the efficacy of the study drug and a significant difference from the placebo group was observed (log-rank test: p=0.0486). The median time to discontinuation due to “inadequate efficacy of the study drug” (time to reach a cumulative discontinuation rate of 50%) was within 150 days in the placebo group, whereas it could not be calculated for the SPP-004 group (final cumulative discontinuation rate>50%).

**Fig 2.**
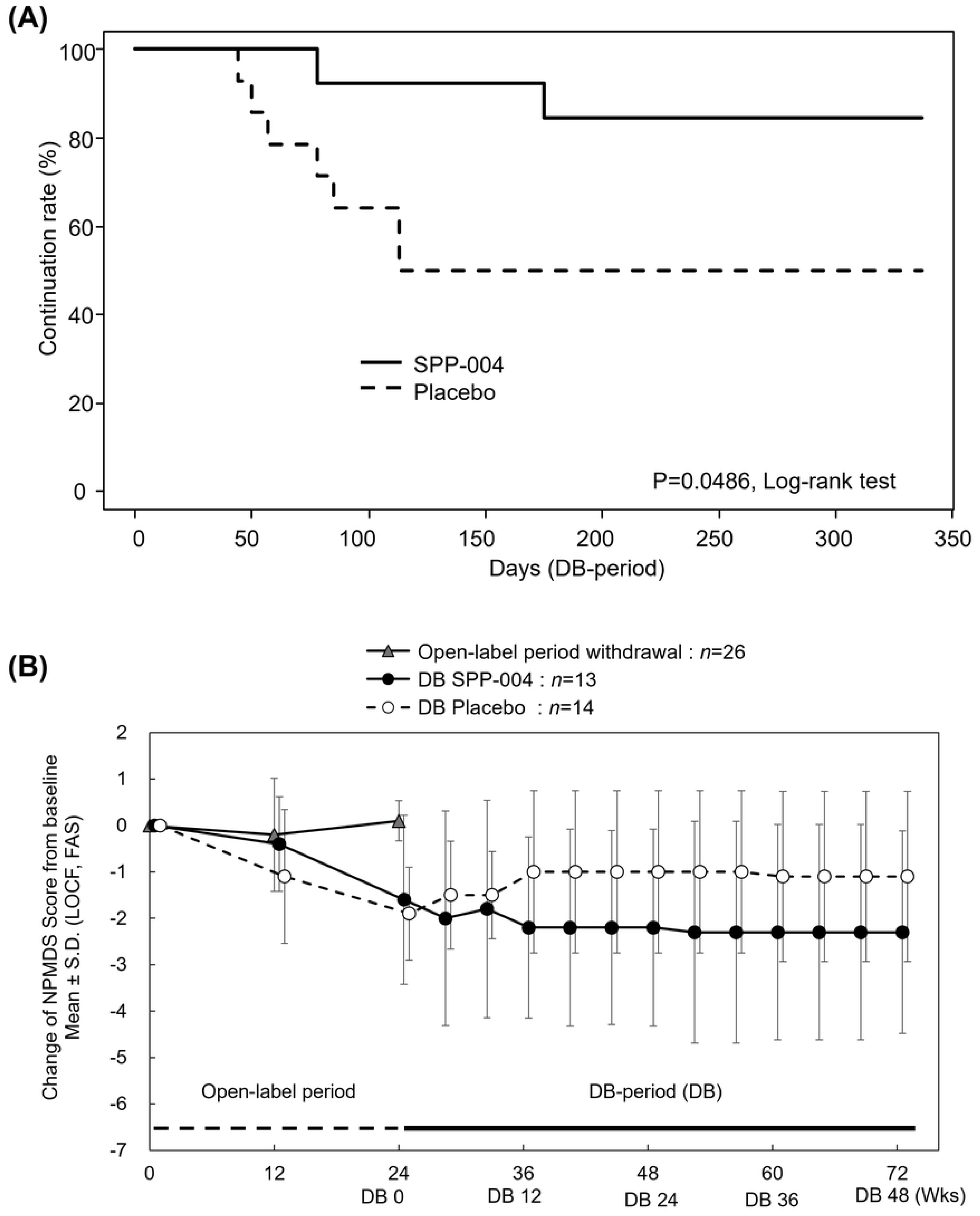
Between-group comparison of changes in efficacy over the study period. (A) Kaplan-Meier plots for the period until discontinuation due to insufficient efficacy, Log-rank test (P<0.05) (B) Change from baseline: NPMDS score (cranial nervous symptoms and myopathy symptoms), mean ±S.D. (LOCF, FAS).

The mean values of changes in the total NPMDS score for cranial nervous symptoms and myopathy symptoms from baseline (FAS, LOCF) were shown in Fig 2B and S2 Table. During the 24-week open-label period, the mean of the NPMDS total scores in the open-label period in the discontinued patients was nearly unchanged (+0.1 ± 0.43). In contrast, the mean values of the changes in the total score during the same period were improved in both the DB-transitioned groups (mean ±S.D.; SPP-004: −1.6 ± 1.82, Placebo: −1.9 ± 1.00). During the 48-week DB-period, the change in score in the placebo group worsened (−1.9 ± 1.00 to −1.1 ± 1.83), while the average total score in the SPP-004 group showed a trend to improve further (−1.6±1.82 to −2.3±2.18).

Fig 3 demonstrates the Swimmer’s plot of each patient during the DB-period, showing maintenance/discontinuation of the effect and improvement/worsening of cranial nervous symptoms and myopathy symptoms that were improved during the open-label period. As mentioned above, the number of patients who discontinued treatment due to inadequate efficacy of the study drug was fewer in the SPP-004 group (2 of 13 patients) than in the placebo group (7 of 14 patients). In addition, the period until discontinuation due to inadequate efficacy of the study drug was longer in the SPP-004 group (77 and 174 days) than in the placebo group (43 to 112 days), suggesting that the efficacy was more sustained in the SPP-004 group (S3 Table).

**Fig 3.**
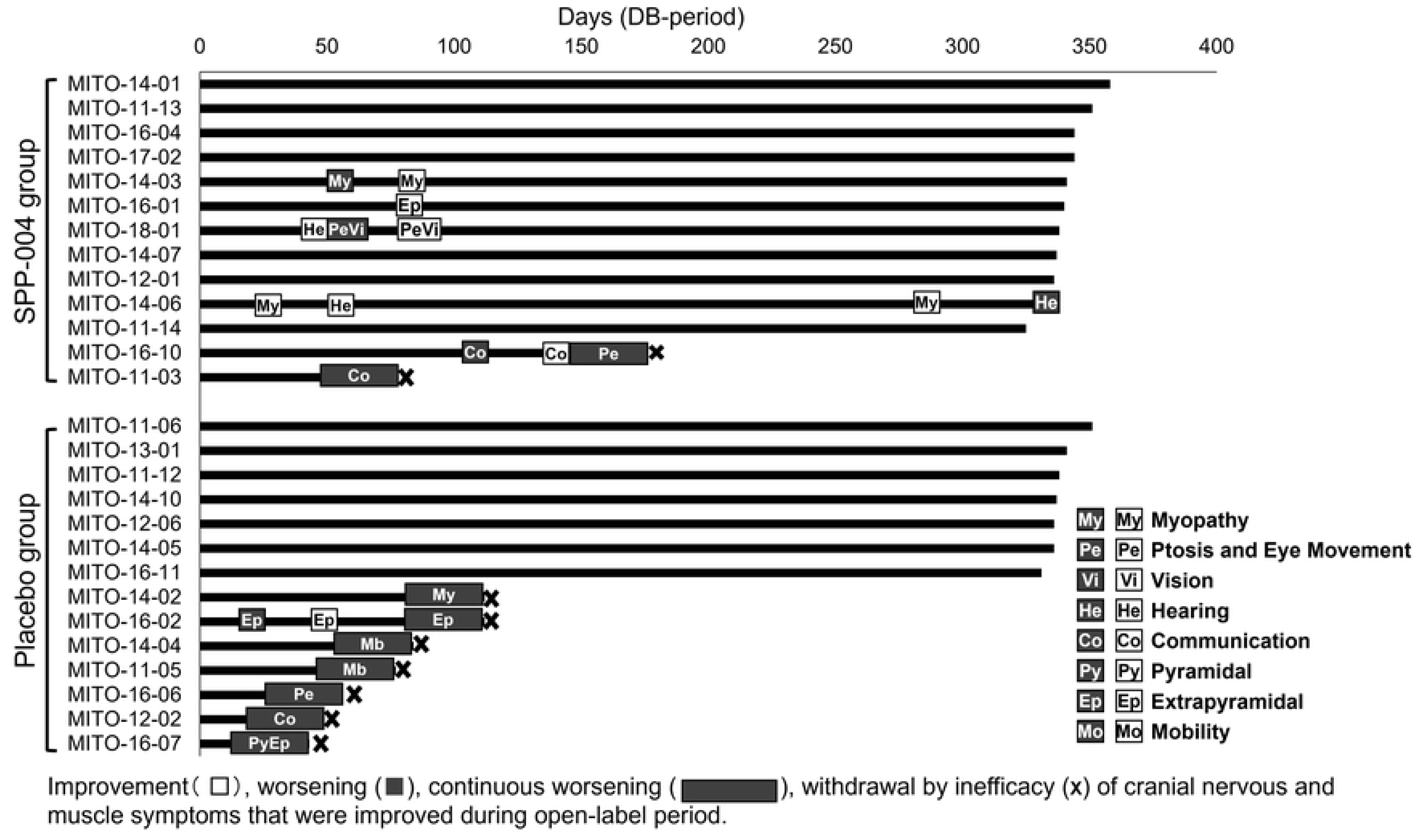
Swimmer’s plot of each patient during the DB-period showing maintenance/discontinuation of the effect and improvement/worsening of cranial nervous symptoms and myopathy symptoms that were improved during the open-label period.

Changes in each item of cranial nervous symptoms and myopathy symptoms from baseline to Week 48 of the DB-period or the time of discontinuation were listed in S4 Table. The efficacy of each item of NPMDS for cranial nervous symptoms and myopathy symptoms was summarized in S5 Table. In the SPP-004 group, efficacy was maintained or improved by 100% except for two items (ptosis and eye movement, communication) during the DB-period. In contrast, in the placebo group, discontinuation due to inadequate efficacy was observed in more than half of the items (6/11 items, mobility, myopathy, pyramidal, extrapyramidal, ptosis and eye movement, and communication) during the same period.

Table 4 summarizes the efficacy evaluated by the number of patients and the number of items with long-term efficacy, short-term efficacy, improvement, and worsening. In terms of the number of patients who showed improvement, there were more patients in the SPP-004 group (6 patients) than in the placebo group (1 patent) during the DB-period (significant difference; p = 0.0329, Fisher’s exact test) (Table 4). In terms of the number of items improved during the open-label period, the numbers were almost the same in both groups, with 13 items in the SPP-004 group and 14 items in the placebo group. However, the number of items that maintained long-term efficacy in the DB-period (SPP-004 group 21 items, placebo group 12 items) and the number of items that improved during the DB-period (SPP-004 group 8 items, placebo group 1 items) were significantly higher in the SPP-004 group (P = 0.0041 and P = 0.0098, respectively, Fisher’s exact test) (Table 4). However, there was worsening during the DB-period in the placebo group (11 items) than in the SPP-004 group (3 items), with a significant difference (p=0.0287, Fisher’s exact test) (Table 4).

**Table 4.**
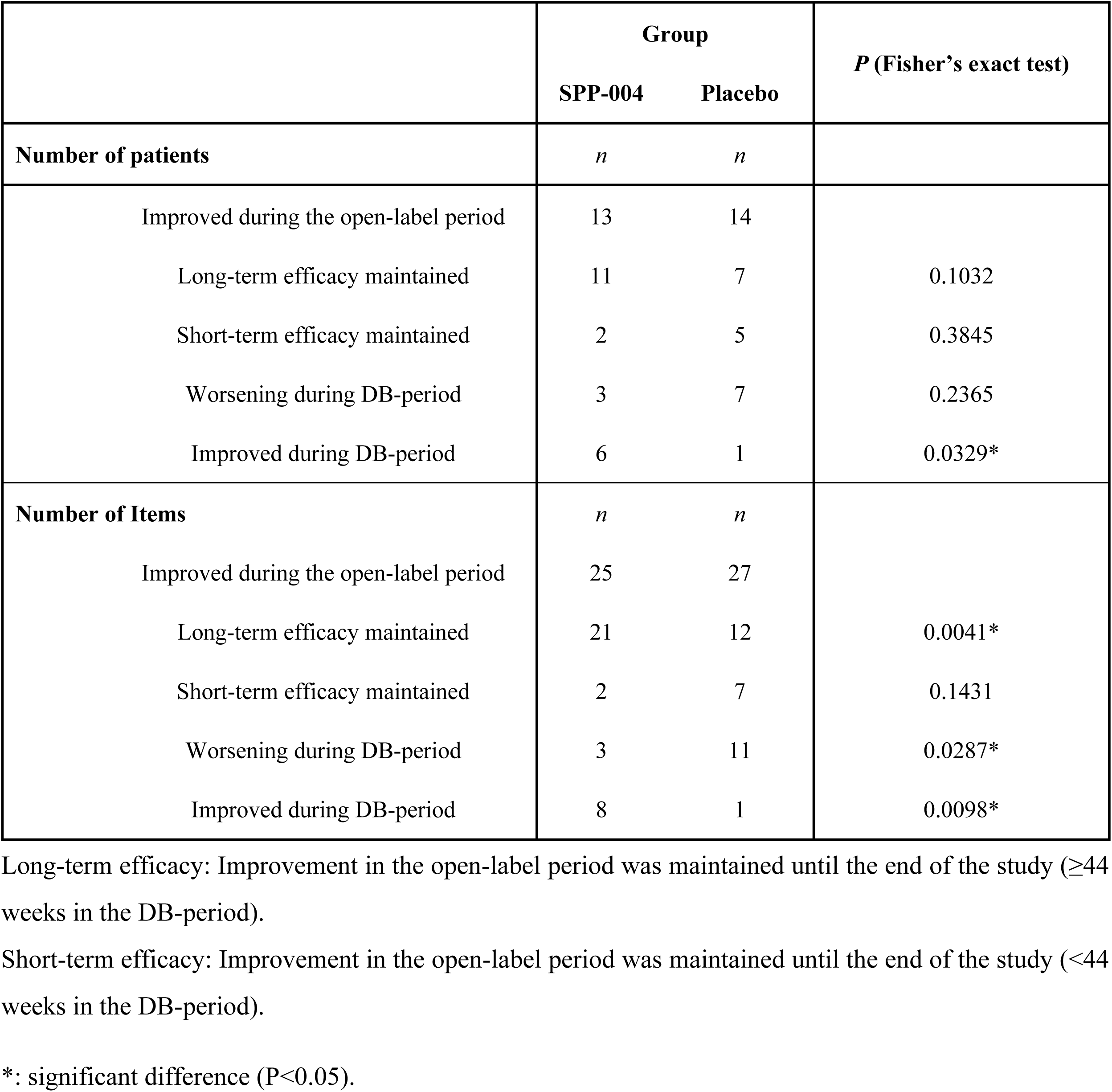
Summary of efficacy evaluated by number of patients and number of items with long-term efficacy, short-term efficacy, improvement, and worsening.

A heat map of changes in scores for each item of cranial nervous symptoms and myopathy symptoms during the whole study period is shown in S1 Fig. Regarding mobility, among 10 patients (SPP-004, 4 patients; Placebo, 6 patients) who improved mobility during the open-label period, the percentage of patients with long-term efficacy was higher in the SPP-004 group (100% [4 out of 4 patients]) than that in the placebo group (66.7% [4 out of 6 patients]). In addition, there were 2 cases that improved only after the transition to the DB-period in the SPP-004 group. However, discontinuation due to inadequate efficacy was observed in the placebo group (33.3% [2 out of 6 patients]).

Regarding myopathy, among 8 patients (SPP-004 3 patients, Placebo 5 patients) who improved myopathy during the open-label period, the percentage of patients with long-term efficacy was higher in the SPP-004 group (100% [3 out of 3 patients]) than that in the placebo group (40.0% [2 out of 5 patients]). Interestingly, 1 patient in the SPP-004 group with a severe symptom (score 3) in myopathy at baseline (MITO-14-06) further improved by 2 points after the transition to the DB-period and turned to be normal (Score 0) after 40 weeks of the DB-period. Though no long-term efficacy was observed in extrapyramidal, the percentage of patients with long-term efficacy of extrapyramidal, ptosis and eye movement, communication, hearing, and neuropathy were also higher in the SPP-004 than in the placebo group (S1 Fig).

Conversely, regarding changes in NPMDS scores in individual patients, 2 patients in the SPP-004 group (MITO-14-06 and MITO-14-07) showed notable improvements in multiple items (S2 Fig). As mentioned above, MITO-14-06 improved the score of myopathy by 3 points, from severe to normal. In addition, hearing also improved by 2 points, from mild to normal from baseline to 8 weeks of the DB-period. However, it worsened at 48 weeks of the DB-period (S2 Fig). Moreover, scores of ataxia and mobility were improved by 1 point after the transition to the DB-period (4 weeks and 16 weeks of the DB-period, respectively). MITO-14-07 improved 5 items (mobility, myopathy, communication, self-care, neuropathy) in the open-label period, and long-term efficacy was maintained for these items. In addition, ptosis and eye movements were improved by 1 point after the transition to the DB-period. A summary of changes in NPMDS scores in these patients is shown in S6 Table.

### Surrogate biomarkers

A heat map of changes in FGF21 and GDF15 in each patient is shown in S3 Fig. At the time of enrolment, there were seven patients with high FGF21 levels (>1,000 pg/ml) and 13 patients with high blood GDF15 (>1,000 pg/ml).

Among the seven patients with high FGF21 levels, three patients, including one patient in the SPP-004 group and two patients in the placebo group, could enter the DB-period. In contrast, four patients failed to enter the DB-period.

Among the 13 patients with high GDF15 levels, six patients, including three patients each in the SPP-004 group and placebo group, could enter the DB-period. In contrast, seven patients failed to enter the DB-period. Among the patients with high GDF15 who entered the period, four patients with moderately high GDF15 (1,000-2,000 pg/ml) maintained the efficacy of the NPMDS score and completed the study period. Conversely, two patients with GDF15 levels over 2,000 pg/ml discontinued at week 12, immediately after entering the DB-period, due to an increase in the total score for NPMDS Sections I and III items 3-8. Among them, one patient with abnormally high GDF15 levels (>4,000 pg/ml) also showed constantly high FGF21 levels (>1,000 pg.ml).

The distribution of blood FGF21 or GDF15 and the NPMDS total score of each patient at the time of enrollment are shown in S4 Fig. Most of the patients had low levels of FGF21 (<300 pg/ml) and GDF15 (<500 pg/ml), which appeared to be not correlated with the total score for NPMDS Sections I and III items 3-8. However, there was a tendency for FGF21 levels to be higher in patients with higher total NPMDS scores. No such tendency was observed for GDF15. However, high (>1000 pg/ml) or extremely high (>4000 pg/ml) levels of GDF15 were restricted to patients with a total NPMDS score of 9 or higher.

Serum lactate levels in the open-label and DB-periods are shown in S5 Fig. At the beginning of the open-label period the mean value of serum lactate exceeded the upper limit of the healthy lactate level (18 mg/ml), and it gradually decreased during the open-label period for 24 weeks (*n*=52). After entering the DB-period, lactate levels increased in the placebo group (*n*=14), while there was a tendency of stabilization of serum lactate level in SPP-004 group (*n*=13) and decreased to the upper limit of healthy lactate level (18 mg/ml) at 48 weeks.

### Safety results

A summary of AEs in the open-label period and DB-period is shown in Table 5. A summary of AEs in the overall study period is shown in S7 Table. The incidence of AEs was 90.7% (49/54) during the overall study period, 87.0% (47/54) during the open-label period, and 92.9% (13/14) in the SPP-004 group and 78.6% (11/14) in the placebo group during the DB-period.

**Table 5.** Summary of Adverse events for the open-label period and the DB-period (SAS)

|  | Open-label period<br>(0-24W) |  | DB-period (0-48W) |  |  |  |  |  |
| --- | --- | --- | --- | --- | --- | --- | --- | --- |
|  | Total<br>*n* =54 |  | SPP-004<br>*n* = 14 |  | Placebo<br>*n* = 14 |  | Total<br>*n* = 28 |  |
|  | Cases (%) | Events | Cases (%) | Events | Cases (%) | Events | Cases (%) | Events |
| All adverse events | 47 (87.0) | 156 | 13 (92.9) | 83 | 11 (78.6) | 46 | 24 (85.7) | 129 |
| Severity |  |  |  |  |  |  |  |  |
| Mild | 26 (48.1) | 109 | 4 (28.6) | 56 | 7 (50.0) | 38 | 11 (39.3) | 94 |
| Moderate | 19 (35.2) | 45 | 7 (50.0) | 24 | 3 (21.4) | 7 | 10 (35.7) | 31 |
| Severe | 2 (3.7) | 2 | 2 (14.3) | 3 | 1 (7.1) | 1 | 3 (10.7) | 4 |
| Causal relationship with<br>investigational drugs |  |  |  |  |  |  |  |  |
| Unrelated | 40 (74.1) | 149 | 13 (92.9) | 83 | 10 (71.4) | 45 | 23 (82.1) | 128 |
| Related | 7 (13.0) | 7 | 0 (0.0) | 0 | 1 (7.1) | 1 | 1 (3.6) | 1 |
| Death | 0 (0.0) | - | 0 (0.0) | - | 0 (0.0) | - | 0 (0.0) |  |
| Serious adverse events | 11 (20.4) | 20 | 9 (64.3) | 20 | 1 (7.1) | 3 | 10 (35.7) | 23 |
| Adverse events leading to<br>discontinuation of study drug | 0 (0.0) | 0 | 1 (7.1) | 1 | 0 (0.0) | 0 | 1 (3.6) | 1 |

Regarding severity, mild was the most common [open-label period: 48.1% (26/54), DB-period: 39.3% (11/28)], followed by moderate [open-label period: 35.2% (19/54), DB-period:35.7% (10/28)], and severe was the least [open-label period: 3.7% (2/54), DB-period: 10.7% (3/28)]. Concerning the causal relationship with the study drug, adverse drug reactions (ADRs) related to the study drug were 13.0% (7/54 patients) during the open-label period and 7.1% (1/14) in the placebo group during the DB-period. There was no ADR in the SPP-004 group during the DB-period. There were no deaths throughout the study period.

The incidence of serious adverse events (SAEs) was 20.4% (11/54) during the open-label period, 64.3% (9/14) in the SPP-004 group, and 7.1% (1/14) in the placebo group during the DB-period (Table 5). The only AE that led to the discontinuation of the study drug was one SAE of oesophageal rupture in the SPP-004 group during the DB-period, for which a causal relationship with the study drug was ruled out.

ADRs classified by SOC and PT are shown in S8 Table. ADRs in the open-label period were tooth discoloration in 4 patients (7.4%) and insomnia, coughing, and vomiting in 1 patient (1.9%), respectively. All events were mild in severity and had a recovery or relief outcome. ADR developed in the DB-period was only one patient (7.1%) with mild diabetes mellitus in the placebo group and not in the SPP-004 group.

The SAEs for the entire period during which SPP-004 was administered are shown in S9 Table. In total, 40 SAEs were developed in 29.6% (16/54) of the patients. The breakdown of SAEs was 6 events of pneumonia in 5 patients (9.3%), 3 events of bronchitis in 3 patients (5.6%), and 1 event in 1 patient each (1.9%) of bacteremia, gastroenteritis, gastroenteritis adenovirus, gastroenteritis viral, influenza, nasopharyngitis, respiratory syncytial virus bronchiolitis, respiratory syncytial virus infection, gastroenteritis norovirus, dehydration, seizure, epilepsy, myoclonus, status epilepticus, upper respiratory tract inflammation, increased upper airway secretion, pleural effusion, pneumonia aspiration, vomiting, oesophageal rupture, and pyrexia, respectively. For all SAEs, a causal relationship to the study drug was ruled out, and the outcome was recovery. Concerning laboratory tests, vital signs, and other observations related to safety, no clinically problematic variations or changes were observed for all patients throughout the study period.

In summary, the safety of SPP-004 was suggested by the following facts that most of the AEs were mild or moderate throughout the period, that all SAEs were denied a causal relationship with the investigational drug, that all ADRs were mild, and that no clinically problematic abnormalities were found in laboratory tests, vital signs, and other observations.

## Discussion

The previous exploratory trial was conducted as a double-blind, placebo-controlled randomized study by enrolling only 10 patients with LS under the age of 24 months [19].

Although the primary observation in the exploratory trial was safety, the double-blinded placebo-controlled study did not sound ethically appropriate for the clinical trial on rare disease infant patients with disease progression. The causal gene abnormalities of LS are highly heterogeneous [5, 6, 33], and the LS symptoms are also very heterogeneous [13]. Therefore, it was difficult to speculate which patients would satisfactorily respond to the supplemental treatment with SPP-004. SPP-004 would improve mitochondrial respiratory activity but could not directly cope with the LS gene abnormalities. The confirmation trial was designed as a double-blind, placebo-controlled, randomized withdrawal trial with patients with LS aged 3 months or older to minimize patient burden. All the patients enrolled were treated with SPP-004 from the beginning, and only the patients who showed improvement in the LS symptoms after the first 24-week treatment were enrolled in the following double-blind, placebo-controlled, randomized withdrawal trial. We could avoid unnecessary prolonged treatments for patients who had little or no benefit from SPP-004 treatment by using this alternative method.

Among 54 patients enrolled in the open-label period, 28 patients showed improvement in some of their typical disease symptoms and were subsequently subjected to the randomized trial to investigate whether their improvements were truly related to SPP-004 treatment. The group of SPP-004 showed continuous improvement except for 2 patients, while half of the placebo group patients got their disease conditions worsened at the early stage after switching SPP-004 to placebo (Fig 3). Interestingly, half of the placebo group maintained their conditions improved until the end of the trial (Fig 3). There are two possibilities to explain the results observed in the placebo group: 1) early treatment was efficacious enough to improve the metabolism in the diseased neuronal and muscular tissues and maintain the improved conditions even after the initial treatment for 24 weeks without further SPP-004 treatment, 2) continuous supplementation of SPP-004 was required to maintain the improved conditions by enhancement of the metabolism in the diseased tissues.

Most AEs were associated with primary diseases, and no AE was biased toward the SPP-004 group, suggesting that twice-a-day administration of SPP-004 for 72 weeks did not differ from placebo administration in terms of safety for patients with LS.

During the DB-period, the cranial nervous and myopathy symptoms were improved in 8 out of 10 patients in the SPP-004 group. Especially 4 types of symptoms, such as mobility, myopathy, pyramidal, and extrapyramidal symptoms, were improved in the SPP-004 group. In contrast, discontinuation was observed in the placebo group due to inadequate efficacy observed in mobility, myopathy, pyramidal and extrapyramidal ptosis, eye movement, and communication. The obvious amelioration of myopathy symptoms was seen in the patient MITO-14-06 in the SPP-004 group (S2 Fig), who had the most severe symptom (score 3: requiring wheelchair use or assistance for mobility, or respiratory compromise) at the time of enrolment, but recovered to a normal level (score 0) by 40 weeks in the DB-period.

The positive effects of 5-ALA and SFC on mobility have been observed in both preclinical and clinical studies [28–32]. Based on those findings and our current results showing improvement in mobility NPMDS, it might be better to focus on the evaluation of the effects on muscular strength and mobile functions.

During the 24 weeks of initial SPP-004 treatment, 28 of 54 patients with LS showed amelioration in 53 symptoms. Among those, 43.4% (23 of 53) had shown an improvement in 12 weeks; however, there was no change observed in eye movement, self-care, and cerebellar ataxia. The average treatment period needed for improvement of NPMDS of cranial nervous and myopathy symptoms was 18.7±6.0 weeks, suggesting that those symptoms were not improved enough during the initial SPP-004 treatment. It might be a reason why we could not observe the remarkable effects of SPP-004 in the previous exploratory trial for only 24 weeks of treatment [19].

Regarding surrogate biomarkers, serum lactate levels indicated positive effects of SPP-004 as reported in our previous study [19]. SPP-004 decreased and stabilized serum lactate levels of the patients in association of stabilization of the total score for NPMDS sections I and III items 3-8, suggesting that serum lactate is a potential biomarker to monitor disease conditions of patients with MD. On the other hand, no good correlations were observed between blood FGF21/GDF15 levels and the total score for NPMDS sections I and III items 3-8. However, there seems to be a tendency for FGF21 levels to increase as NPMDS scores increase. In addition, high (>1000 pg/ml) or extremely high (>4000 pg/ml) levels of GDF15 were limited to patients with a total MPMDS score of 9 or higher. Therefore, these blood protein markers weakly reflect the status of symptoms of mitochondrial diseases.

The limitation of this confirmation trial was partly due to the unavailability of information on LS-associated genetic alterations identified in each patient. Over 75 genes in mitochondrial and nuclear DNA have been identified as a cause of LS, and the symptoms and prognosis depend on the abnormality of the causal genes [33,34]. Shimura et al. [15] reported that skin fibroblasts harvested from six out of eight patients with MD including LS responded to 5-ALA treatment and improved oxidative phosphorylation and increased ATP biosynthesis in mitochondria. In contrast, two of them did not show any response to 5-ALA. Considering those facts, we may speculate a relationship between the causal gene abnormality and the severity of LS, as well as the response to SPP-004. Additional investigation of the gene alterations in each patient with LS may help to identify patients who could benefit from SPP-004 treatment.

The doses of SPP-004 were determined by the body weight classification based on the data obtained in the exploratory trial (SPED-ALA-001). The doses used in the previous trial were calculated based on the safety and tolerability data of 5-ALA-HCl and SFC in Phase I with healthy adult volunteers of Caucasian and Japanese (SBI Pharmaceutical, In-house document), and on the *in vitro* mitochondrial respiration activity data using fibroblasts harvested from MD patients [15]. Dose optimization may be reevaluated by using an appropriate LS mouse model if available.

Nevertheless, considering the observations in the present confirmation trial, we conclude that SPP-004 had some benefits to the patients who could enter the double-blind placebo-controlled randomized trial after the initial treatment with SPP-004 for 24 weeks. Those patients were nearly half of the patients with LS enrolled in the confirmation study. Lim et al. [6] reported that the probability of severe disease burden (NPMDS>25) was higher in the patients with disease onset at <6 months than at >6 months. In the previous study, the average of the total score for NPMDS sections I and III items 3-8 in the patients with early-onset (<6 months) was higher (i.e., severe symptoms) than that in the patients with late-onset (>6 months), suggesting that early disease onset may be associated with poor prognosis and severe symptoms. Therefore, the treatment with SPP-004 needs to start as early as possible after the disease onset.

The supplements composed of 5-ALA-phosphate and SFC as health authority-approved food ingredients have been commercially available in several countries and areas of the world, including Japan, Bahrain, Abu Dhabi, Jordan, Makao, and Singapore. Various physiological and psychological benefits of 5-ALA-phosphate and SFC in a healthcare supplement formulation have been reported in Japan, Bahrain, and USA as follows: 1) anti-diabetic effects [35–38], 2) improvement of sleep quality [39], 3) improvement of mood and coping ability [40], 4) enhancement of respiratory efficiency and training performance in older women [30,31], 5) improvement of depressive mentality in middle-aged women [32], 6) improvement of symptoms of late-onset hypogonadism in aged men [41], 7) improvement of language ability in an ATR-X syndrome patient, one of the X-linked intellectual disability syndromes caused by mutations in the *ATRX* gene, characterized by male patients, severe intellectual disability, α-thalassemia (HbH), and autistic behavior [42], 8) improvement of sarcopenia symptoms [43], 9) improvement of COVID-19 symptoms and sequelae [44,45].

Some of those clinical study results are theoretically supported by the findings obtained in the *in vitro* and *in vivo* preclinical studies by using Zucker diabetic fatty rats [46], *ALAS-1* heterozygous mice [47], Alzheimer model mice [15], ATRX model mice [48], autism model rats [16], sarcopenia model mice [49], low-density lipoprotein receptor knockout mice [50] and SARS-CoV-2 infected cells [51,52].

Considering progressive nature of LS [6], the stabilization or prevention of progression by treatment with SPP-004 may be expected. The supplemental use of healthcare nutrition composed of 5-ALA-phosphate and SFC in the same molecular ratio as that of 5-ALA-HCl and SFC in SPP-004 might be applicable for the treatment of patients with MD under the MD therapy guidelines.

We conclude that SPP-004, composed of 5-ALA-HCl and SFC, was safe in patients with LS, and its therapeutic effects on the symptoms of nearly half of patients with LS enrolled were observed. SPP-004 may be beneficial to patients with progressive LS by stabilizing or improving disease conditions when it is given at the early stage of LS after the disease onset.

## Data Availability

All relevant data are within the manuscript and its Supporting Information files.

## Data availability

The datasets generated during and/or analyzed during the current study are available from the corresponding author upon reasonable request.

## Ethics statement

The clinical trial was conducted in accordance with the World Medical Association Declaration of Helsinki, the Japanese Act on Securing Quality, Effectiveness, and Safety of Pharmaceuticals and Medical Devices, the Ministerial Ordinance on Standards for Conducting Clinical Trials of Pharmaceuticals, and Good Clinical Practice. Approval for this study was mainly granted by the ethics committee of Saitama Medical University Hospital (approval code number 977), and the protocol was approved by the ethics committees at each participating institute. The written consent of a patient’s legal representative (or guardian) was obtained before the enrollment of patients in the trial.

## Author contributions

Conceptualization: AO YA KM KT MY MN

Data curation: AO YA KM HS HO SK (Satoko Kumada) TI HK YH

Formal analysis: AO YA KM HN MN

Funding acquisition: AO MN

Investigation: AO YA KM HS HO SK (Satoko Kumada) TI HK YH SA MA YU KN KI AM HK YN

Methodology: AO YA KM HN KT MY MN

Project administration: AO YA KM KF KT MY MN

Resources: AO MY MN

Software: AO YA HN

Supervision: AO MN

Validation: YA HN SK (Shunichi Kurose) KF

Visualization: HN MN

Writing – original draft: HN MN

Writing – review & editing: AO YA KM HS HO SK (Satoko Kumada) TI HK YH SA MA YU KN KI AM HK YN HN SK (Shunichi Kurose) KF KT MY MN

## Acknowledgments

The authors are thankful to Fumiaki Kobayashi and Kazuo Nakamura, CTD, for their coordination of the trial and their helpful discussions on the protocol preparation and interpretation of the results. We are grateful to Ikuma Musha, Hiroshi Kawana, and Yutaka Ueda at Saitama Medical University, Shinji Minowa and his colleagues in Clinical Trial Office of Saitama Medical University Hospital for their coordinating this clinical trial. We also highly appreciate all the members of SBI Pharmaceuticals who dedicated their efforts to initiate and complete this trial through management of finance, test drug production, quality assurance, safety and regulatory affairs.

## Supporting information

**S1 Table. Dosage and administration of Investigational drug.**

**S2 Table. Change from baseline: NPMDS score (cranial nervous symptoms and myopathy symptoms), mean ±S.D. (LOCF, FAS).**

**S3 Table. Period until discontinuation due to inadequate efficacy of study drug.**

**S4 Table. Changes in cranial nervous symptoms and myopathy symptoms in each patient (baseline to DB-period 48 weeks or at the time of discontinuation).**

**S5 Table. Summary of efficacy of each item of cranial nervous symptoms and myopathy symptoms in the DB-period.**

**S6 Table. Summary of changes in NPMDS scores for cranial nervous symptoms and myopathy symptoms in notably improved patients (MITO-16-06 and MITO-14-07). The improvement ratio indicates the percentage of the number of improved items to the number of symptomatic items at baseline.**

**S7 Table. Summary of adverse events (overall study period*)**

**S8 Table. Adverse drug reaction for the open-label period and the DB-period (SAS).**

**S9 Table. Serious adverse events (overall study period*)**

**S1 Fig. Heatmap diagram showing changes of each score of cranial nervous symptoms and myopathy symptoms in each patient (patients with no change in score of the item were excluded).**

**S2 Fig. Heatmap diagram showing changes in score of each item in the notably improved patients.**

**S3 Fig. Heatmap diagram showing changes in FGF21 and GDF15 in each patient.**

**S4 Fig. Distribution of blood FGF21 or GDF15 and NPMDS total score for each patient at the time of enrollment.**

**S5 Fig. Serum lactate levels in open-label period and DB period.**

